# Successful classification of clinical pediatric leukemia genetic subtypes via structural variant detection using HiFi long-read sequencing

**DOI:** 10.1101/2024.11.05.24316078

**Authors:** Lisa A. Lansdon, Byunggil Yoo, Ayse Keskus, Irina Pushel, Chengpeng Bi, Tanveer Ahmad, Asher Bryant, Adam Walter, Margaret Gibson, Mary Rindler, Weijie Li, Sultan M. Habeebu, Linda D. Cooley, John Herriges, Elena Repnikova, Lei Zhang, Keith J. August, Terrie G. Flatt, Alan S. Gamis, Erin M. Guest, J. Allyson Hays, Maxine Hetherington, Karen Lewing, Tomi Pastinen, Mikhail Kolmogorov, Midhat S. Farooqi

## Abstract

Gene fusions are common primary drivers of pediatric leukemias and are the result of underlying structural variant (SVs). Current clinical workflows to detect such alterations rely on a multimodal approach, which often increases analysis time and overall cost of testing. In this study, we used long-read sequencing (lrSeq) as a proof-of-concept to determine whether clinically relevant (cr) SVs could be detected within a small (n = 17) pediatric leukemia cohort. We show that this methodology successfully determined all known crSVs detected through routine clinical testing. We also identified crSVs, such as an ins(11;10)(q23.3;p12p12) forming a *KMT2A*::*MLLT10* fusion, missed by routine clinical approaches, resulting in the classification of leukemia genetic subtypes for four additional patients. This study demonstrates the diagnostic potential of lrSeq as an assay for SV detection in pediatric leukemia and supports lrSeq as a valuable tool for the accurate detection of crSVs.

## Introduction

Pediatric leukemias are the most common cancers of childhood; they are largely defined by genetically distinct subgroups that confer prognostic significance [1-5]. Gene fusions are common genetic drivers of many of these subgroups and often result from underlying chromosomal translocations and/or other structural variants (SVs) [1, 3, 6, 7]. Current routine clinical diagnostic testing algorithms for pediatric leukemia rely on a series of complementary yet distinct approaches to identify these SVs, including karyotyping, fluorescence *in situ* hybridization (FISH), chromosomal microarray, DNA sequencing, RNA sequencing (followed by expression profiling and/or fusion panel analysis), and, finally, optical genome mapping: an emerging technology in clinical laboratory practice [6, 8-11]. Although generally effective, this approach has several drawbacks: it necessitates that sufficient neoplastic sample be available to perform a battery of tests until a genetic driver alteration is identified; it is often limited to neoplastic-only (also known as “tumor-only”) analyses, which cannot distinguish between true somatic events versus those that are in the patient’s germline; and, finally, it results in an overall increased cost of testing and time to diagnosis due to the use of multiple different methods.

Long-read sequencing (lrSeq) allows for the improved resolution of historically difficult-to-sequence regions of the genome as well as superior detection and delineation of SVs compared with other existing technologies [12, 13]. This approach has been used for variant detection in rare disease [14, 15], and other studies have demonstrated the utility of lrSeq in adult cancer testing, which successfully revealed novel SVs in colorectal cancer [16], lung cancer [17], mantle cell lymphoma [18], acute myeloid leukemia (AML) [19], and hereditary cancer risk [20, 21], among others [22, 23]. Some lrSeq studies have been performed in a pediatric cancer setting, notably for pediatric medulloblastoma [24]. However, the diagnostic utility of lrSeq for pediatric cancer, and especially for pediatric leukemia, has not been systematically evaluated: one study has demonstrated successful SV detection from lrSeq data in a pediatric B-ALL cell line [25], but no patient cohort studies have been published to date.

Here, we investigate the feasibility of SV detection by HiFi lrSeq in a small pediatric leukemia cohort (n=17 cases) selected from patients with known (n=5) and unknown (n=12) genetic subtypes. Patients from historically under-sequenced populations (i.e., of Hispanic/Latino or multiracial ethnicities) were prioritized for this study. The demographics of the cohort assessed in this study are summarized in Figure 1 (see also Supplemental Table 1). The lrSeq data generated from archived DNA for each case were assessed using a recently developed somatic SV detection tool, Severus [26], which uses a phased breakpoint graph approach for the resolution of complex rearrangements [27-30]. Severus can both utilize paired tumor/normal data for improved, true somatic SV detection, or assess neoplastic data in a “tumor-only” fashion (i.e., without a paired normal sample). It also showed the best performance compared to currently available lrSeq SV callers when assessing a panel of tumor cell lines [26]. We first aimed to determine whether lrSeq paired with SV calling by Severus in both tumor/normal and tumor-only modes could result in the accurate detection of SVs in cases with known alterations. We then explored whether additional cases with unknown genetic subtypes after routine clinical testing might be resolved by this approach.

**Figure 1:**
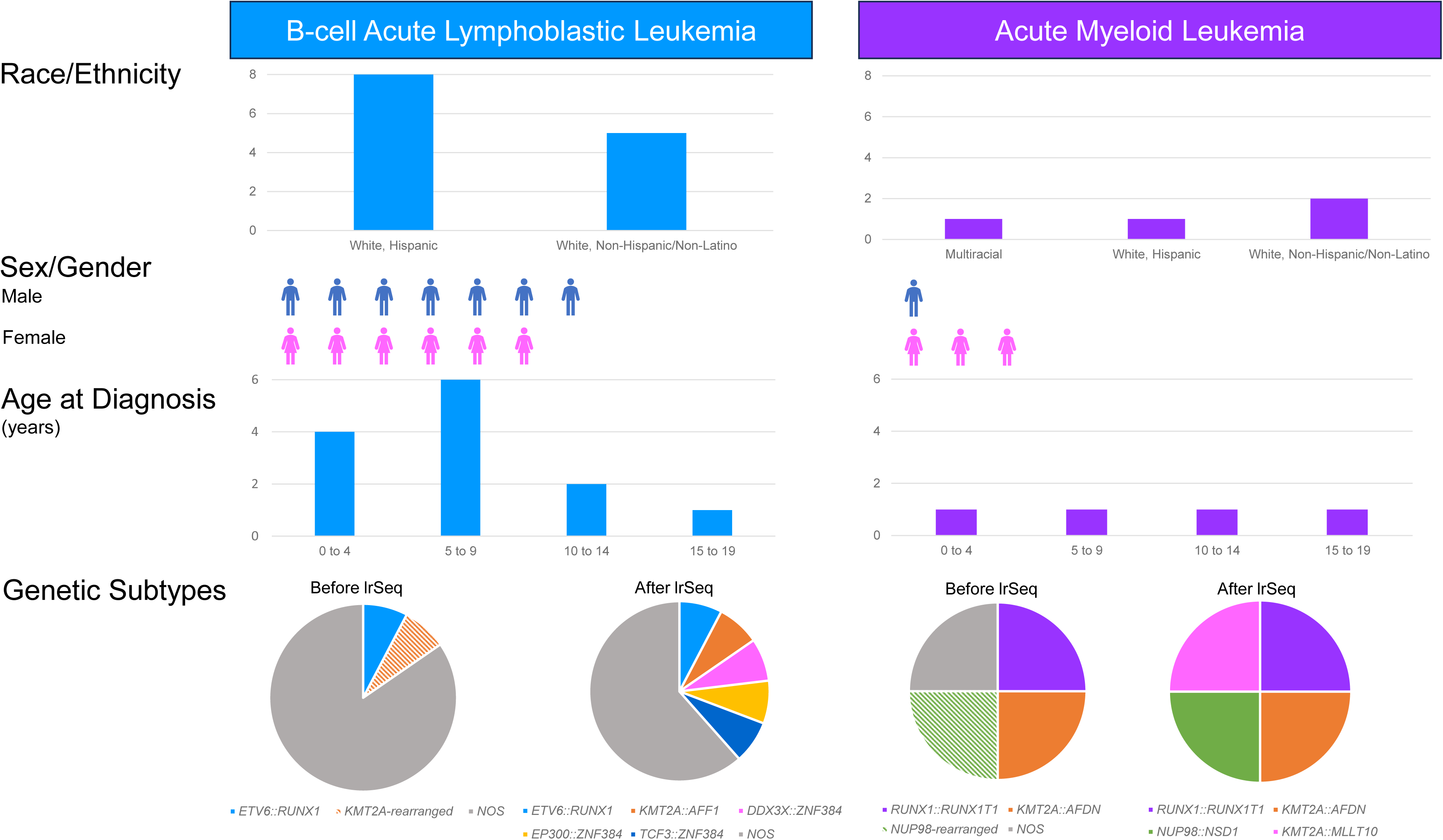
Pediatric Leukemia Cohort Demographics. 17 pediatric patients were included in the present study, comprising of 13 individuals with B-cell acute lymphoblastic leukemia and 4 individuals with acute myeloid leukemia. The race/ethnicity, sex/gender, and age at diagnosis for each patient are specified. In addition, genetic subtypes of the 5 “known” and 12 “unknown” genetic subtypes (including the 4 “unknown” cases that were resolved in the present study) are depicted. Striped pie chart segments indicate cases for which one of the two partner genes involved in the structural variant was not known prior to lrSeq.

## Results

### Tumor/Normal Analysis of Leukemia Samples with Known Structural Variants

Five tumor/normal pairs had known, diagnostically significant SVs previously detected by routine clinical testing. All five of these cases were successfully detected by lrSeq and Severus. This included two B-ALLs, one with a t(12;21)(p13;q22) resulting in *ETV6*::*RUNX1* fusion and the other with a t(4;11)(q21;q23) expected to result in *KMT2A::AFF1* fusion (formerly known as *MLL::AF4*), and three AMLs, one with t(8;21)(q22;q22) resulting in *RUNX1::RUNX1T1* fusion, one with t(6;11)(q27;q23) resulting in *KMT2A::AFDN* fusion, and one with a *NUP98* rearrangement identified via FISH but without additional testing performed to identify the partner gene. Breakend analysis of the lrSeq data using Severus detected each of these rearrangements. In addition, Severus provided the exact genomic breakpoints within each gene, confirmed that *AFF1* was rearranged with *KMT2A* in the t(4;11)(q21;q23) B-ALL, and also identified the unknown fusion partner in the AML case with the *NUP98* rearrangement, calling a t(5;11)(q35;p15.5) resulting in *NUP98::NSD1* fusion. The specific details of these rearrangements, including the breakpoints within each gene, are listed in Supplemental Table 1.

### Tumor/Normal Analysis of Genetically Undefined Leukemia Samples

We next examined four tumor/normal pairs with a heretofore genetically undefined leukemia subtype. These four cases had previously undergone routine clinical genetic characterization but did not have a WHO diagnostic genetic subtype identified. All four had lrSeq DNA data analyzed for clinically relevant SVs. On average, Severus detected an average of 14 somatic breakend calls per sample (range 0-60; see Supplemental Table 2), and two of these four genetically unknown cases were successfully defined by this approach (see Supplemental Table 1). One was an AML with an ins(11;10)(q23.3;p12p12) resulting in *KMT2A::MLLT10* fusion (Figure 2), and the other was a B-ALL with a t(12;19)(p13;p13) resulting in *TCF3::ZNF384* fusion (Figure 3).

**Figure 2:**
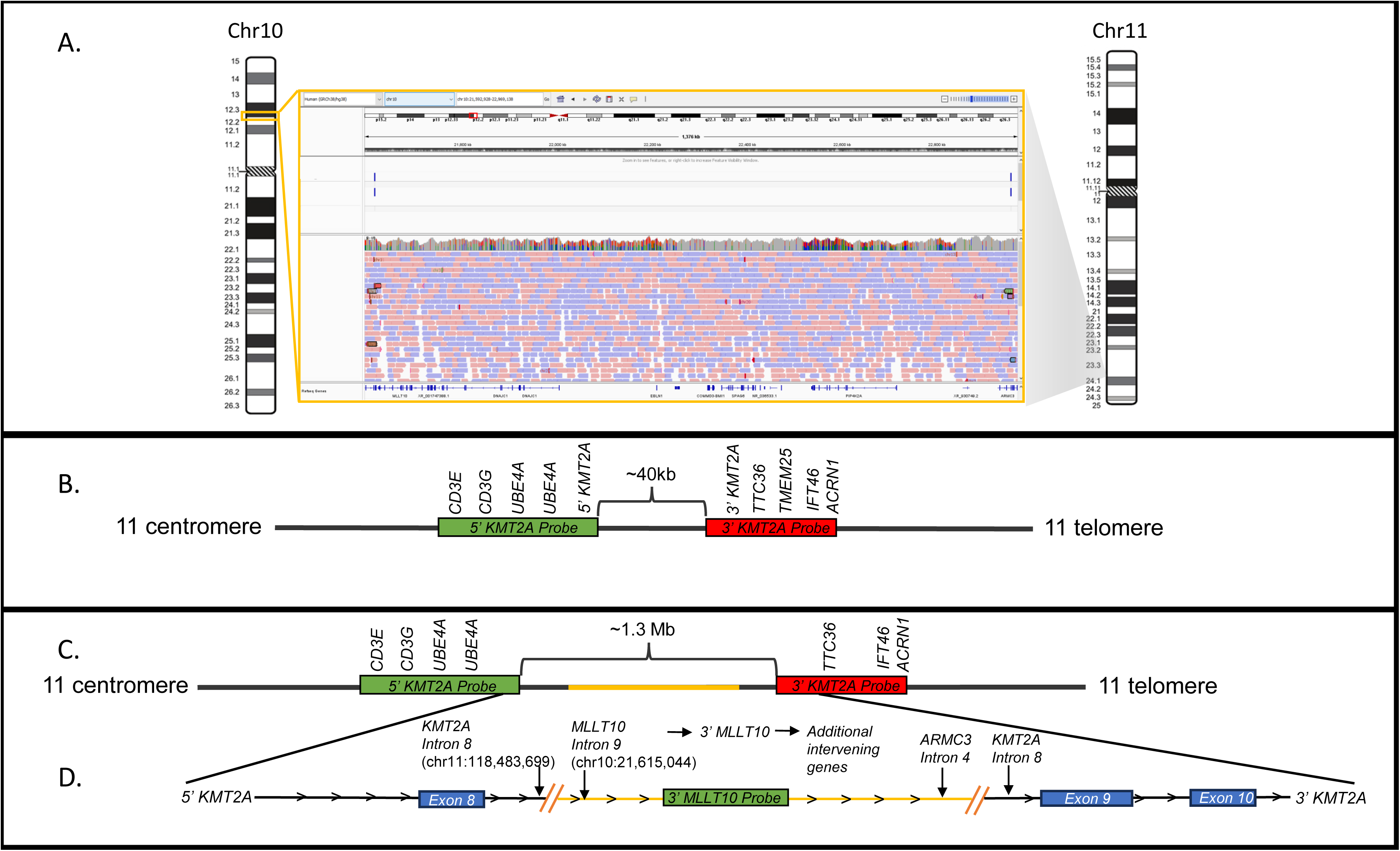
KMT2A::MLLT10 fusion detected by Severus in a previously genetically undefined acute myeloid leukemia case. A) Detection of an ∼1.3 Mb segment of 10p12 that is called by Severus (indicated by the blue bars in the haplotagged_severus track of the IGV screen capture) and corresponding long-read sequencing (lrSeq) reads. This segment is inserted into the *KMT2A* gene on chromosome 11q23. Note: the red and blue coloring of the reads does not denote pair orientation (e.g., +/-) in a lrSeq setting; it is provided here only for visual ease. B) Typical spacing of the *KMT2A* breakapart FISH probe utilized for the in-house acute myeloid leukemia (AML) panel. The probes are represented by their respective colors. C) Illustration of the increased spacing of the probes that is expected after the insertion of the ∼1.3 Mb segment of 10p12 (indicated by the yellow bar) into *KMT2A*. D) Schematic of the orientation of the *KMT2A* (NM_001197104.2) and *MLLT10* (NM_001195626.3) genes including the intronic location of each breakpoint; a functional fusion product is predicted.

**Figure 3:**
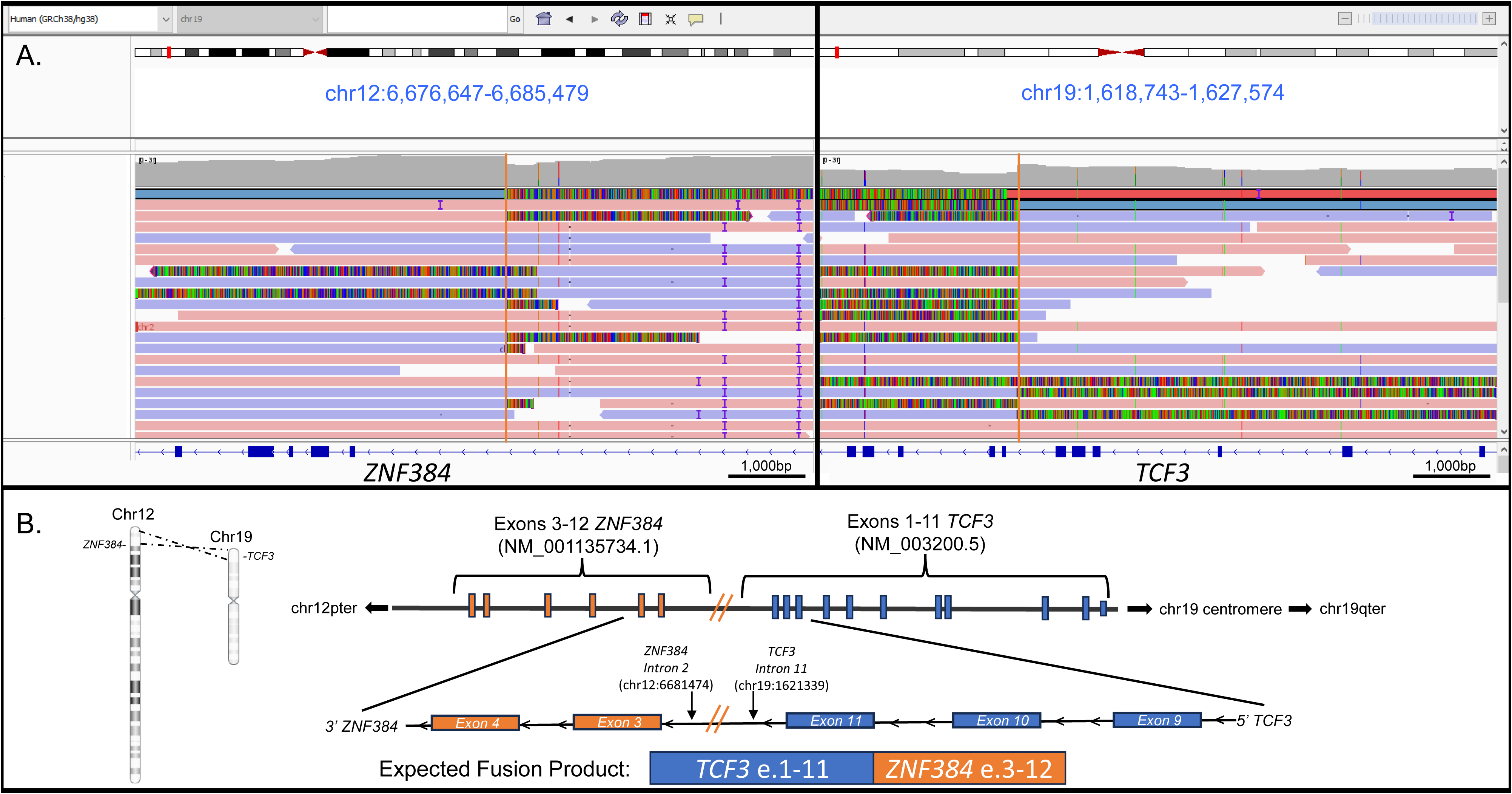
t(12;19)(p13;p13) detected by Severus in a previously genetically undefined precursor B-cell acute lymphoblastic leukemia case. A) IGV demonstrating the presence of chimeric reads, indicating a translocation between *TCF3* and *ZNF384*. B) Schematic of the orientation of *TCF3* (NM_003200.5) and *ZNF384* (NM_001385745.1) genes including the intronic location of each breakpoint. Note that a functional fusion product is predicted. Note: the red and blue coloring of the reads does not denote pair orientation (e.g., +/-) in a lrSeq setting; it is provided here only for visual ease.

The AML with the ins(11;10)(q23.3;p12p12) was further investigated to compare lrSeq findings with those of prior clinical genetic testing, especially since a FISH AML panel (which included a *KMT2A* breakapart probe) had been performed clinically but was negative. Manual review of the lrSeq data demonstrated that an ∼1.3 Mb segment of chromosome 10p within 10p12.33p12.2 (extending from intron 9 of *MLLT10* (NM_001195626.3) to intron 4 of *ARMC3* (NM_173081.5)) was inserted into intron 8 of the *KMT2A* gene (NM_001197104.2) on 11q23.3 (Figure 2A). This insertion would cause the 5’ and 3’ probes of *KMT2A* to further separate from their baseline of ∼40kb (i.e., no rearrangement present; Figure 2B), but only to a total distance of ∼1.4Mb: a difference too small to visualize in FISH or chromosome analyses (i.e., cytogenetically cryptic; Figure 2C). Follow-up testing using a breakapart *MLLT10* FISH probe confirmed the presence of a balanced rearrangement involving *MLLT10* (Supplemental Figure 1). Furthermore, the use of lrSeq data allowed for the exact breakpoints of this balanced insertion to be assessed, which demonstrated that a functional fusion product would be expected due to the 5’ to 3’ orientation of the inserted 10p sequence (Figure 2D). Of note, RNA was not available for this case for additional confirmatory testing.

The SV calls from the B-ALL with the t(12;19)(p13;p13) were similarly compared with prior clinical testing results to try and resolve the discrepancy in SV detection. Analysis of the SV breakpoints demonstrated that this was a balanced reciprocal translocation between chromosomes 12p13.31 and 19p13.3 (Figure 3A, 3B), with a presumed functional product resulting from the fusion of the 5’ end of *TCF3* (NM_003200.5) through intron 11 with intron 2 of *ZNF384* (NM_001385745.1) through the 3’ end of the gene on the derivative chromosome 19 (Figure 3C). Given its cytogenetically cryptic and balanced nature, this SV was not detected by the prior clinical chromosome and array studies, and RNA-based fusion testing had not been performed for this case. Clinical FISH testing (an ALL panel) had been done, but neither of the genes involved in the SV (*ZNF384* or *TCF3*) were tested. These genes are commonly absent from standard clinical laboratory ALL FISH panels, since translocations involving these genes are infrequently observed in pediatric B-ALL [31]. Although *TCF3* is now a part of our clinical ALL FISH panel, a *ZNF384* FISH probe has not yet been validated.

### In silico Assessment of Limit of Detection

We next explored the ability of lrSeq and Severus to detect SVs across a range of tumor amounts. Notably, all neoplastic samples utilized for this study had high blast percentages at diagnosis (∼80-94% blasts per Pathology review). Thus, *in silico* modeling was performed to determine the limit of detection (LOD) for this approach at an average of 30x depth of coverage (which corresponds to using 1 HiFi sequencing SMRT cell per sample). Reads from the nine tumor samples with paired normal data were ‘diluted’ using sequence data that had been generated from the normal sample (see Supplemental Table 3 for a representative example). This analysis demonstrated that the lower limit of detection was ∼3/30 reads, which corresponded with a 1:10 dilution (i.e., LOD of a 10% VAF in a sample with ∼30x coverage) and suggests a minimum required blast or tumor percentage of ∼20% per neoplastic sample for clonal SV detection.

### Assessment of Tumor-Only Structural Variant Calling

Although paired tumor/normal analysis enables true somatic SV calling from lrSeq data, obtaining a germline sample within a clinically relevant timeframe is not always feasible in pediatric leukemia testing, nor are paired ‘normal’ samples always available for research cohorts. Thus, we ran Severus in “tumor-only mode” (i.e., without the paired normal sample) for the previously analyzed tumor/normal pairs above (n=9). There was an expected increase in the average number of cluster IDs (consisting of breakend calls that comprise unique SVs) in the unmatched samples versus the matched dataset (Supplemental Figure 2; see Supplemental Tables 4 and 5 for the resulting breakend call files from the matched tumor/normal versus tumor-only analysis, respectively). On average, Severus called an average of 285 breakend calls per case (range 124-440). However, when restricting to breakends occurring within a gene, the number of calls was reduced by approximately half (see Supplemental Table 2 for counts per case by calling mode). To further screen these calls, we employed a list of genes known to be mutated in pediatric leukemia per the National Cancer Institute’s Childhood Cancer Genomics summary and/or the World Health Organization’s diagnostic genetic subtypes of B-ALL and AML [32, 33]. This approach readily identified the crSVs in all 7 of 7 neoplastic samples (100%) from the tumor/normal analyses above (see Supplemental Table 6 for the gene lists utilized), resulting in an average of 1 breakend call per case (range 0-4; see Supplemental Table 2). This suggests that the use of appropriate filters in a tumor-only dataset is equally effective as paired tumor/normal testing despite the larger number of SVs initially detected. Of note, a few ongoing studies are aiming to generate population-scale SV databases using lrSeq data, which could be used as a panel of normal in tumor-only sample analysis in the future [34, 35].

To further investigate the utility of tumor-only lrSeq and analysis using Severus, eight additional B-ALL cases without an available germline sample, and also without a known genetic subtype after routine clinical testing, were analyzed. By using the filtering approach outlined above, a crSV was identified in two of these eight cases: one t(X;12)(p11.4;p13.31) resulting in *DDX3X::ZNF384* fusion; and one t(12;22)(p13.31; q13.2) resulting in *EP300::ZNF384* fusion. A summary of all fusions identified by Severus in both the “known” and “unknown” cases analyzed can be found in Figure 1 (with breakpoints of each SV listed in Supplemental Table 1). As before, the existing clinical data for both of these ‘tumor-only, unknown’ cases were reviewed to determine why these rearrangements had not been previously detected.

Both cases had received karyotyping, FISH B-ALL panel, and chromosomal microarray testing. However, *ZNF384* rearrangements are commonly cytogenetically cryptic [36, 37] and thus were not detected by chromosomes. Because ZNF384 probes are not part of standard clinical B-ALL FISH panels, *ZNF384* was not investigated by FISH for either case. Finally, the *EP300::ZNF384* rearrangement was genomically balanced and thus not detected by microarray. For the other case (*DDX3X::ZNF384*), the microarray analysis showed significant genomic complexity in the 12p region, where *ZNF384* resides, including several copy number alterations which made the genetic interpretation difficult. No additional testing was performed for the case with the *DDX3X::ZNF384* rearrangement, and RNA was not available for follow-up testing. Interestingly, the case with the *EP300::ZNF384* fusion did receive RNA-based fusion analysis testing as well as next-generation sequencing (NGS). However, *ZNF384* was not included on the RNA fusion panel utilized for the analysis, nor was the NGS assay validated for SV detection. Thus, neither rearrangement was detected via the clinical test algorithm selected for each case.

### lrRNA-seq Fusion Detection Supports the crSVs Identified by lrSeq

In addition to lrSeq of DNA for SV detection, three cases underwent lrRNA sequencing (lrRNA-seq) to assess the feasibility of this approach for fusion transcript detection as well as to determine whether the crSVs detected by Severus were supported by the presence of RNA fusion transcripts. Diagnostically relevant fusion transcripts were successfully detected in 2/3 cases, which demonstrated a *KMT2A::AFDN* and an *EP300::ZNF384* fusion product; these transcripts resulted in the fusion of exon 11 of *KMT2A* (NM_001197104.2) with exon 2 of *AFDN* (NM_001386888.1), and exon 7 of *EP300* (NM_001429.4) with exon 1 of *ZNF384* (NM_001385745.1), respectively (Figure 4). The *KMT2A::AFF1* transcript that was expected in the other B-ALL case was not detected by the fusion caller and also was not visible upon manual inspection. *KMT2A* fusion transcripts within the context of infant B-ALL (as in the present case) may be expressed at low levels and therefore difficult to detect [38]. Thus, the inability to detect the *KMT2A::AFF1* transcript in the present analysis suggests that technical optimization of lrRNA-seq by our laboratory (e.g., attempting concatenation of cDNA to increase lrRNA-seq transcript capture [39]) is still needed to detect all clinically relevant fusions. Overall, the successful identification of fusion transcripts in two cases supporting the crSVs and their corresponding DNA breakpoints highlights the clinical utility of lrSeq in pediatric leukemia crSV detection.

**Figure 4:**
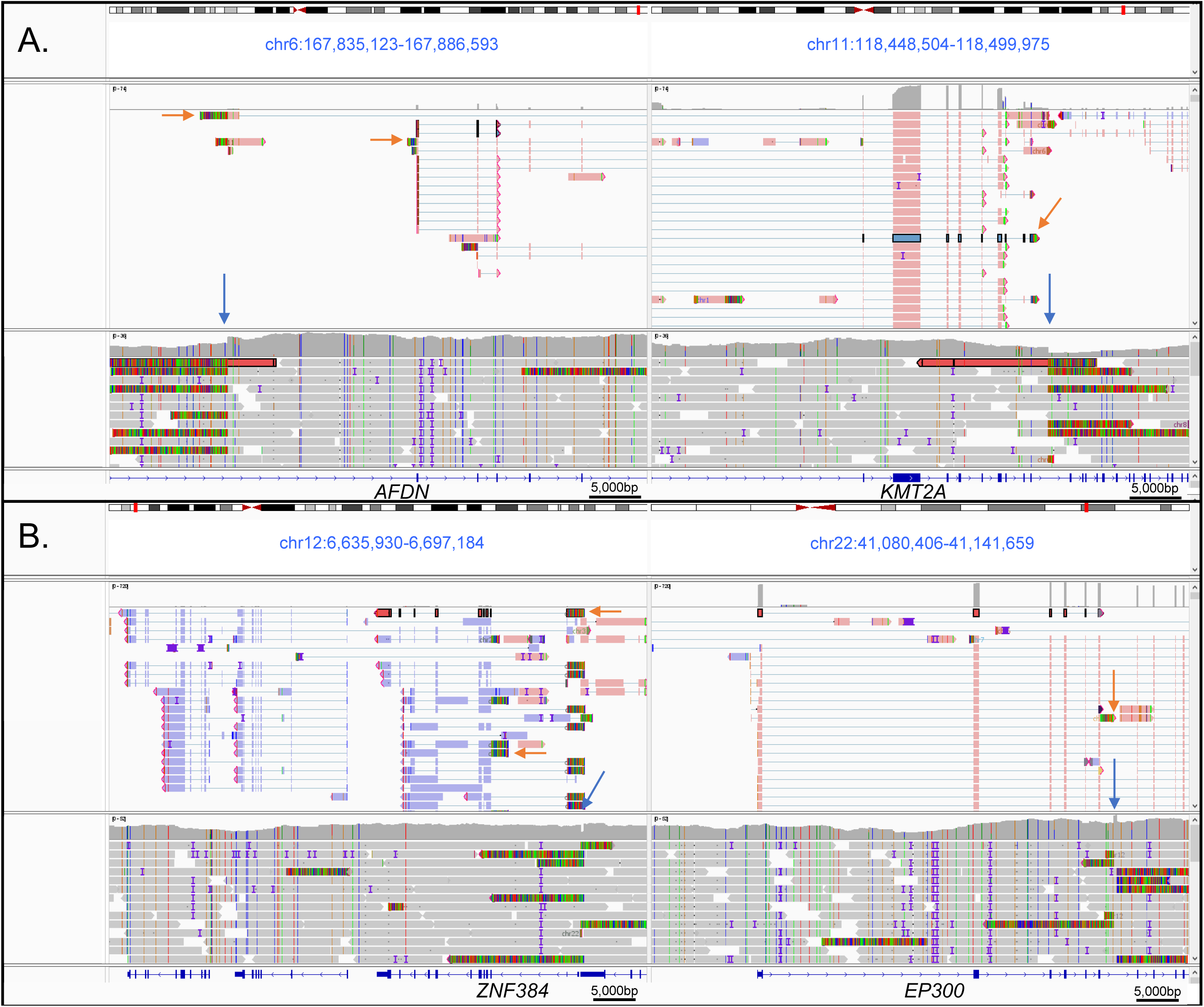
lrRNA-seq successfully detects fusion transcripts within pediatric leukemia cases. A) IGV demonstrating the presence of a *KMT2A::AFDN* fusion in the lrRNA-seq data (top panel) which corresponds with the lrSeq DNA breakpoints (bottom panel). B) IGV demonstrating the presence of an *EP300::ZNF384* in the lrRNA-seq data (top panel) which corresponds with the lrSeq DNA breakpoints (bottom panel). Blue arrows indicate the SV breakpoints as called by Severus in the lrSeq DNA data. Orange arrows indicate representative fusion transcripts present within the screen capture. Of note, various isoforms can be observed for the *KMT2A::AFDN* fusion case (A).

## Discussion

Long-read sequencing (lrSeq) is an effective technology for structural variant (SV) detection in inherited and neoplastic settings, especially for SVs in repetitive regions of the genome or those that are of high genetic complexity [13, 19, 40-42]. Although several studies have assessed SV calling using lrSeq in various types of adult cancer [16-23], the potential to use it as a clinical diagnostic tool for patients with cancer has not been evaluated systematically. In addition, extremely limited focus has been given to pediatric cancer, with only one published study to date investigating the utility of SV detection using lrSeq in pediatric medulloblastoma [24]. Furthermore, these studies were performed using existing bioinformatics SV callers, which had primarily been built for germline analyses. Severus (a bioinformatics tool specifically optimized to detect somatic SVs using lrSeq data) has only recently been developed [26]. Since Severus outperformed other long-read somatic SV detection tools [26], this study was performed to assess the feasibility of detecting diagnostically relevant SVs using breakend calling from lrSeq data in a small clinical pediatric leukemia cohort.

Using this approach, all diagnostically relevant somatic SVs were identified with the exact breakpoints demonstrated in the five patients who had a genetic classification of their leukemia known through prior clinical diagnostic testing. Of note, for two of these five cases, lrSeq resolved the partner gene, which enhanced the prior clinical results. In addition, lrSeq and Severus analysis resolved the genetic driver alterations for four cases (4/12 total unknowns) that had not been genetically classified by prior clinical testing. Thus, in this small exploratory cohort, the present approach resulted in a 33% increase in diagnostic yield over current clinical diagnostic methodologies. It is important to note, however, that the clinical utility of RNA-based profiling with fusion transcript detection and Optical Genome Mapping (OGM) [10, 43, 44] are of known benefit for genetically subtyping pediatric leukemia [8, 9], but these assays have historically not been a routine part of our clinical testing workflow. Had they been, it may have aided in the detection of underlying structural variants that were cryptic by other methodologies. However, both approaches present their unique challenges.

For one, RNA is not always available, especially for historical cases, and requires specific isolation and storage conditions. It is also a challenge to maintain optimal conditions during transport for such specimens if they are being shipped out to external reference laboratories for testing. Additionally, for clinical laboratories that employ RNA-based fusion panels, if one of the genes involved in the fusion is not specifically included on the panel, then that fusion transcript will be missed. Such is the case for *ZNF384*, which is not part of the gene list for multiple clinically available RNA-based hematologic cancer fusion panels, even at Children’s Oncology Group (COG) approved laboratories or those at large pediatric academic medical centers.

For OGM, although it is a DNA-based technology, it presently requires a specific isolation procedure for ultra-long DNA molecules, which is not always compatible with banked samples. In contrast, the lrSeq data analyzed in our study were generated using DNA that was isolated using routine clinical protocols (see STAR Methods). Additionally, the breakpoint resolution of SVs detected via OGM can be pinpointed only to a genomic interval that is a few kilobases in length as it is dependent upon the location of the breakend relative to the repeat sequences that OGM examines. For comparison, lrSeq allows for the unbiased and successful detection of the exact nucleotide-level breakpoints of each rearrangement. Thus, although the diagnostic yield of our small cohort would likely be impacted if RNA profiling, fusion analysis, and/or OGM was incorporated into our clinical workflow, this study demonstrates a proof-of-principle for DNA-based lrSeq to be successfully, and perhaps more feasibly, implemented clinically for SV detection in pediatric leukemia.

Importantly, the successful detection of these SVs provides highly relevant information that has a significant and direct impact on patient management. This is indeed the case for the four genetically ‘unknown’ cases above that were solved by lrSeq and Severus. For example, the AML patient with the ins(11;10)(q23.3;p12p12) identified in the present study would have been managed as high-risk, whereas patients without a diagnostic genetic alteration are managed as intermediate risk. Patients that are high-risk undergo more aggressive treatment, such as hematopoietic stem cell transplantation [45] and *KMT2A*-rearranged AML has been shown to respond more favorably to gemtuzumab ozogamicin [46], which would have been an additional therapy that could have been pursued for this patient.

Similarly, *ZNF384* rearrangements, like the three identified in the previously ‘unknown’ cohort, are known to be karyotypically cryptic within the context of pediatric B-ALL, requiring detection via *ZNF384* FISH testing and/or next-generation DNA or RNA sequencing. *ZNF384* is also known to rearrange with one of several partner genes, most commonly *EP300*, *TCF3*, *TAF15*, and *CREBBP* [47]. Identifying the fusion partner is important in *ZNF384*-rearranged B-ALL as *EP300* confers an inferior prognosis and high-risk disease [47]. In contrast, the partner genes *TCF3* or *DDX3X* are not expected to be of prognostic significance, but the identification of a *TCF3::ZNF384* or *DDX3X::ZNF384* translocation helps rule out the presence of a different, previously undetected, high-risk genetic alteration.

The utility of lrRNA-seq for isoform detection and inferring structural variants has been described previously in pediatric oncology settings [25, 48]; however, it has not yet been investigated within individuals with pediatric leukemia. Therefore, we were interested in determining whether diagnostically relevant fusion transcripts supporting the structural variants detected via DNA-based lrSeq and Severus could be captured by this approach. Both the *KMT2A::AFDN* and *EP300::ZNF384* fusion isoforms were successfully detected, suggesting that this test could be feasibly implemented as an orthogonal method to further support findings from DNA-based SV detection and/or as a replacement for short-read RNA-seq. Furthermore, lrRNA-seq may additionally complement DNA-based SV detection by allowing for the identification of genomic rearrangement-independent fusions [49]. As mentioned above, the third case that underwent lrRNA-seq was unsuccessful at detecting the known *KMT2A::AFF1* fusion. However, this was not entirely surprising; although *KMT2A* rearrangements are relatively reliably detected via a variety of methodologies (e.g., FISH and chromosomes), *KMT2A* fusions may be expressed at low levels in some cases of infant B-ALL [38]. Thus, increased depth of coverage (thereby improving RNA transcript capture) and/or an optimized approach to specifically detect fusions in this setting would be beneficial. Finally, lrRNA-seq offers increased resolution of fusion transcripts in that it accurately captures specific isoforms; this study demonstrated that these isoforms matched the DNA-based lrSeq breakpoints with remarkably high precision (as shown in Figure 4). Future work will be needed to determine the most clinically relevant isoform(s) for each patient as lrRNA-seq becomes more widely implemented.

In summary, this study establishes the successful detection of crSVs by lrSeq in a small pediatric leukemia cohort, including an increased diagnostic yield over commonly used clinical workflows. Given that lrSeq can generate complete genomic data for a sample in a few days and its cost has decreased to less than $1,000 USD per genome, we may be reaching a point where its clinical implementation in cancer, especially in a pediatric setting, is now feasible. Finally, since lrSeq provides additional genetic information (such as the ability to detect single nucleotide variants, copy number variants, aneuploidy, methylation profiling, etc.) not investigated in the present study, it is likely that the utility of lrSeq as a comprehensive cancer profiling test will continue to grow.

## Data Availability

All data produced in the present study are available upon reasonable request to the authors. Sequence data will be deposited in a repository for controlled access upon manuscript publication.

## Acknowledgements

M.S.F. and E.G. thank Braden’s Hope for Childhood Cancer, Elizabeth and Monte McDowell, the Black & Veatch Foundation, and Big Slick for their generous support, and Nhu Bui and Mindy Spano for their assistance with financial administration. M.S.F., E.G., and L.A.L. also thank Children’s Mercy Oncology Biorepository study personnel: Judy Vun, Amie Hatfield, and Robin Ryan; as well as Jason Seymour and Keiondra Sanders in the Children’s Mercy Research Institute (CMRI) Biorepository, for their assistance with sample collection and processing. M.S.F. and L.A.L. are grateful to Jonas Korlach for his expertise and thoughtful feedback on the manuscript, and to Jonathan Bibliowicz, Gary Gelland, Khi Pin Chua, Aaron Wenger, and Alex Sockell for their collaborative support. A.K., T.A., A.B and M.K. were supported in part by the Intramural Research Program of the NIH. PacBio provided partial reagent support for this study.

## Author contributions

MSF and LAL conceptualized this analysis. LAL, BY, AK, MK, and MSF analyzed the data. LAL, BY, IP, MK, and MSF drafted the paper. All authors provided edits for the final submission.

## Declaration of interests

The authors declare no competing interests.

## Supplemental information

Supplemental Table 1: *Demographic information of the study participants and diagnostic SVs identified via lrSeq.* \**TCF3* was not part of the standard FISH ALL panel at the time of diagnosis for this patient. TN = paired tumor/normal run mode; TO = tumor-only run mode.

Supplemental Table 2: *Breakend counts per sample by calling mode and filters utilized*

Supplemental Table 3: *In-silico modeling to determine the SV limit of detection for tumor/normal pairs*

Supplemental Table 4: *Severus breakend calls using a paired tumor/normal approach*

Supplemental Table 5: *Severus breakend calls using a tumor-only approach*

Supplemental Table 6: *Pediatric leukemia genes utilized for Severus breakend analyses*

Supplemental Figure 1: *FISH and chromosome analyses for the cryptic KMT2A::MLLT10 AML case.* The boxes in the karyotype indicate the chromosomes where *MLLT10* and *KMT2A* reside; both chromosome pairs look normal. *KMT2A* breakapart FISH does not demonstrate any visible probe separation. However, the *MLLT10* breakapart FISH probe demonstrates that a translocation is present.

Supplemental Figure 2: *Assessment of unique cluster IDs called by Severus in tumor/normal vs tumor-only mode*.

Breakend counts were determined by using unique cluster IDs generated by Severus for each patient when assessing the neoplastic (“tumor”) sample with the matched normal sample (blue), in the absence of the matched normal sample (“tumor-only”; orange), or in the absence of the matched normal and filtering by breakends occurring within genes (green).

## STAR Methods

### Lead Contact

Further information and requests for resources and reagents should be directed to and will be fulfilled by the lead contact, Midhat S. Farooqi.

### Materials Availability

This study did not generate new unique reagents.

### Data and Code Availability

HiFi DNA-seq data will be deposited at dbGAP and are publicly available as of the date of publication. Accession numbers will also be listed in the key resources table. This paper does not report original code. Any additional information required to reanalyze the data reported in this paper is available from the lead contact upon request.

### Study Participant Details

Nine pediatric leukemia cases with corresponding normal (remission) samples were selected for HiFi sequencing from the Children’s Mercy Research Institute Biorepository (CRIB) following informed consent. These cases were enrolled in the CRIB between 2017 and 2023 and had sufficient material available for HiFi sequencing. The pathology diagnosis for five of these cases was precursor B-cell acute lymphoblastic leukemia (B-ALL), and for the remaining four was acute myeloid leukemia (AML). Five of the cases (2 B-ALL; 3 AML) had diagnostic translocations that had been identified clinically using routine cytogenetic testing (FISH, chromosomes, and microarray). The remaining four cases (3 B-ALL; 1 AML) had undergone routine clinical testing (FISH, chromosomes, microarray, next-generation sequencing panel or exome sequencing), and were selected because no diagnostic genetic driver alteration (i.e., genetic driver “not otherwise specified”; NOS) had been identified. In addition, eight B-ALL cases without known genetic drivers and for which a paired normal sample were not available were selected based upon their NOS genetic status, and samples that were available from historically under-sequenced populations were prioritized. The age, sex, gender, race, and ethnicity of each patient are provided in Supplemental Table 1, when provided by self or physician report. The ancestry and socioeconomic status of the study participants were not captured when patients enrolled in the CRIB study.

### Method Details

Genomic DNA from each sample (tumor and normal) was extracted using either a Chemagen (PerkinElmer, Baesweiler, Germany) in accordance with each manufacturer’s protocol or via manual TKM isolation. DNA samples were mechanically fragmented by pipet action on the Diagenode Megaruptor 3, and library preparation was performed with the SMRTbell Prep Kit 3.0 (Pacific Biosciences (“PacBio”), Menlo Park, CA, USA). Size selection for intact library fragments greater than 10kb was performed using the PippinHT (Sage Science, Beverly, MA, USA), and fragment size and distribution was confirmed using a Femto Pulse bioanalyzer (Agilent, Santa Clara, CA, USA). The concentration of each library was determined by Qubit, and then libraries were sequenced on a PacBio Revio instrument using supported reagents (Revio Polymerase Kit, Revio Sequencing Plate and Revio SMRT Cell Tray) to a target depth of 30x. The PacBio Human Whole Genome Sequencing (WGS) workflow was used to process HiFi reads for haplotagged alignment and phasing, followed by structural variant calling from each tumor / normal (TN) pair using Severus [26]. The TN Severus output was filtered for breakend calls and assessed for overlap with genes known to be relevant to pediatric leukemia (see Supplemental Table 6). All prior clinical genetic testing used for SV detection for each case was collated and used for comparisons with the breakend analysis findings (Supplemental Table 1). Each neoplastic sample was additionally processed by Severus without the paired normal sample to assess the feasibility of diagnostic SV detection in a tumor-only (TO) setting. The TO Severus output was similarly filtered for breakend calls and analyzed for overlap with genes known to be relevant to pediatric leukemia, then compared to the TN Severus output in addition to the existing clinical genetic data that was available for each case.

For lrRNA-seq RNA was isolated from cell pellets in TRIzol (ThermoFisher, Cat. No. 15596026) using a TRIzol/chloroform precipitation and were cleaned with a RNeasy Mini Kit (Qiagen, Cat. No. 74104). The samples were then quantified with a Qubit RNA BR Assay Kit (ThermoFisher, Q10210) to determine the concentration of the sample. Samples with a high concentration (>800ng/µL) were diluted 1:10 with RNase-free water and were quantified again. The 1:10 diluted samples were then checked on a RNA ScreenTape (Agilent, Cat. No. 5067-5577 and 5067-5576) on the TapeStation platform to assess the RIN score prior to Iso-Seq library preparation to ensure that the RIN was greater than or equal to 7.0. Long-transcript cDNA libraries were prepared following the procedure described in the Iso-Seq™ Express Template Preparation for Revio Systems protocol from Pacific Biosciences. The maximum recommended input (300ng) of RNA was aliquoted from the 1:10 diluted RNA samples and was used as input in the protocol. The samples were enriched for long transcripts greater than 3kb in length. Quantification with a Qubit dsDNA HS Assay Kit (ThermoFisher, Cat. No. Q32854) determined that the cDNA yield of the samples was less than the minimum required yield (160ng) for the Revio system and that the samples would need additional PCR cycling as described in Appendix 1 of the Iso-Seq procedure. 3 additional PCR cycles were performed on the samples, as recommended by the yields table listed in Appendix 1, with NEBNext High-Fidelity 2X PCR Master Mix (NEB, Cat. No. M0541S) used in the PCR reamplification master mix. The samples were then bead-cleaned following the procedure for low-yielded samples and were quantified again with a Qubit dsDNA HS Assay Kit to ensure there was enough cDNA for SMRTbell library preparation.

Up to 500ng of cDNA from each sample was used as an input into SMRTbell library preparation without pooling using the SMRTbell Express Template Prep Kit 2.0 (Pacific Biosciences, 100-938-900). After the SMRTbell library cleanup, the libraries were quantified with a Qubit dsDNA HS Assay Kit to determine their final concentration. The library size was determined with a High Sensitivity D5000 ScreenTape (Agilent, Cat. No. 5067-5592 and 5067-5593) on the TapeStation platform. Resulting libraries were sequenced with one Revio SMRT Cell (Pacific Biosciences, 102-202-200) each on the Revio System using the Revio Polymerase Kit (Pacific Biosciences, 102-739-100) according to standard manufacturer’s directions with loading at 90pM per library.

### Key Resources Table

| REAGENT or RESOURCE | SOURCE | IDENTIFIER |
| --- | --- | --- |
| Deposited data |  |  |
| HiFi sequencing data | dbGAP | Will be added prior to publication. |
| Software and algorithms |  |  |
| PacBio WGS Workflow | GitHub | https://github.com/PacificBiosciences/HiFi-human-WGS-WDL/commit/f5de5444dc1f2aaa24f194b29540ba73a511302b |
| Severus | GitHub | https://github.com/KolmogorovLab/Severus |

## Notes

### Competing Interest Statement

The authors have declared no competing interest.

### Funding Statement

This study was funded by Braden's Hope for Childhood Cancer, Elizabeth and Monte McDowell, the Black & Veatch Foundation, and Big Slick.
A.K., T.A., A.B, and M.K. were supported in part by the Intramural Research Program of the NIH.
PacBio provided partial reagent support for this study.

### Author Declarations

The IRB of Children's Mercy Kansas City gave approval for this work.

