## Supplementary figures and images for "Successful classification of clinical pediatric leukemia genetic subtypes via structural variant detection using HiFi long-read sequencing"

### Supplemental Figure 1

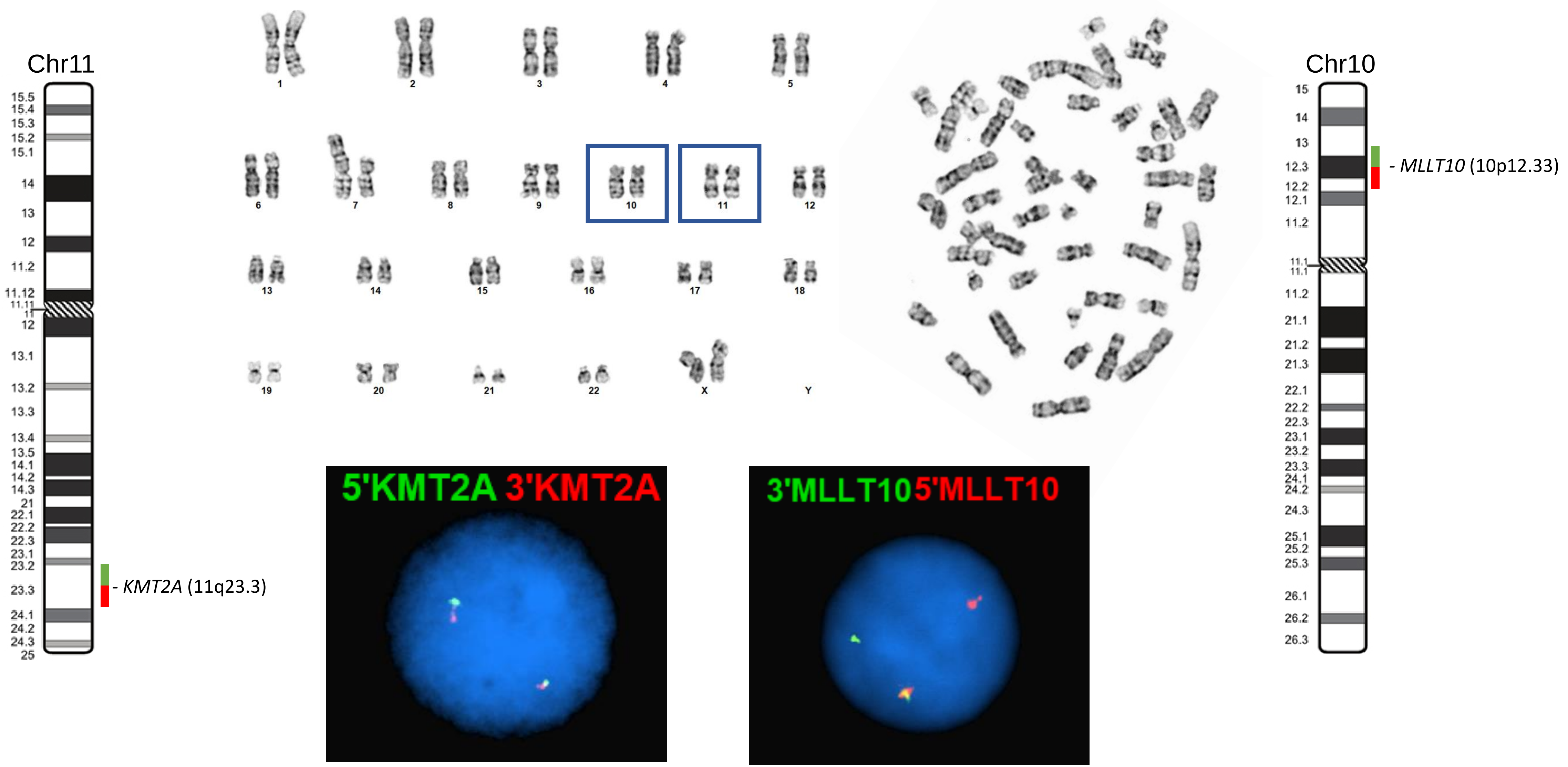

### Supplemental Figure 2

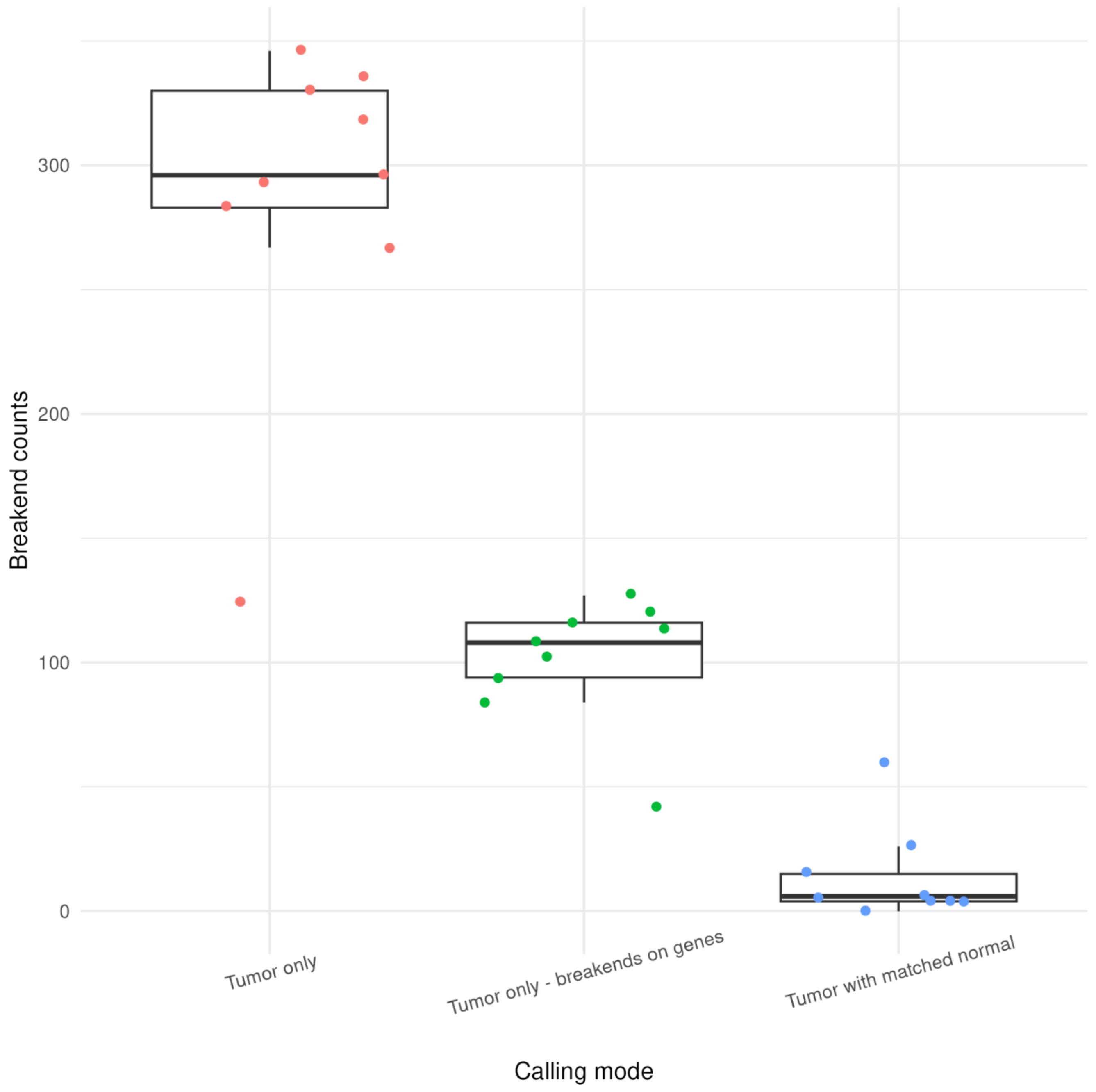
