## Supplemental Table 3 for "Successful classification of clinical pediatric leukemia genetic subtypes via structural variant detection using HiFi long-read sequencing"

| **Sample** | **Pct tumor** | **Cluster ID** | **SV Type** | **Distance to nearest gene** | **Gene ID** | **Gene Name** | **Chr1** | **Chr1 Coord** | **Chr2** | **Chr2 Coord** | **Strands** | **Supp. Read** |
| --- | --- | --- | --- | --- | --- | --- | --- | --- | --- | --- | --- | --- |
| Case 4 | 1 | severus_19 | BND | 0 | ENSG00000119772 | *DNMT3A* | chr2 | 25286660 | chr4 | 87078149 | ++ | 16 |
| Case 4 | 1 | severus_20 | BND | 0 | ENSG00000119772 | *DNMT3A* | chr2 | 25286681 | chr11 | 118488607 | -- | 11 |
| Case 4 | 1 | severus_19 | BND | 0 | ENSG00000172493 | *AFF1* | chr4 | 87078149 | chr2 | 25286660 | ++ | 16 |
| Case 4 | 1 | severus_21 | BND | 0 | ENSG00000172493 | *AFF1* | chr4 | 87078190 | chr11 | 118488445 | -+ | 16 |
| Case 4 | 1 | severus_21 | BND | 0 | ENSG00000118058 | *KMT2A* | chr11 | 118488445 | chr4 | 87078190 | -+ | 16 |
| Case 4 | 1 | severus_20 | BND | 0 | ENSG00000118058 | *KMT2A* | chr11 | 118488607 | chr2 | 25286681 | -- | 11 |
| Case 4 | 0.5 | severus_8 | BND | 0 | ENSG00000119772 | *DNMT3A* | chr2 | 25286660 | chr4 | 87078149 | ++ | 8 |
| Case 4 | 0.5 | severus_9 | BND | 0 | ENSG00000119772 | *DNMT3A* | chr2 | 25286681 | chr11 | 118488607 | -- | 6 |
| Case 4 | 0.5 | severus_8 | BND | 0 | ENSG00000172493 | *AFF1* | chr4 | 87078149 | chr2 | 25286660 | ++ | 8 |
| Case 4 | 0.5 | severus_10 | BND | 0 | ENSG00000172493 | *AFF1* | chr4 | 87078190 | chr11 | 118488445 | -+ | 9 |
| Case 4 | 0.5 | severus_10 | BND | 0 | ENSG00000118058 | *KMT2A* | chr11 | 118488445 | chr4 | 87078190 | -+ | 9 |
| Case 4 | 0.5 | severus_9 | BND | 0 | ENSG00000118058 | *KMT2A* | chr11 | 118488607 | chr2 | 25286681 | -- | 6 |
| Case 4 | 0.25 | severus_3 | BND | 0 | ENSG00000119772 | *DNMT3A* | chr2 | 25286660 | chr4 | 87078149 | ++ | 4 |
| Case 4 | 0.25 | severus_4 | BND | 0 | ENSG00000119772 | *DNMT3A* | chr2 | 25286681 | chr11 | 118488607 | -- | 4 |
| Case 4 | 0.25 | severus_3 | BND | 0 | ENSG00000172493 | *AFF1* | chr4 | 87078149 | chr2 | 25286660 | ++ | 4 |
| Case 4 | 0.25 | severus_5 | BND | 0 | ENSG00000172493 | *AFF1* | chr4 | 87078190 | chr11 | 118488445 | -+ | 5 |
| Case 4 | 0.25 | severus_5 | BND | 0 | ENSG00000118058 | *KMT2A* | chr11 | 118488445 | chr4 | 87078190 | -+ | 5 |
| Case 4 | 0.25 | severus_4 | BND | 0 | ENSG00000118058 | *KMT2A* | chr11 | 118488607 | chr2 | 25286681 | -- | 4 |
| Case 4 | 0.2 | severus_3 | BND | 0 | ENSG00000119772 | *DNMT3A* | chr2 | 25286660 | chr4 | 87078149 | ++ | 4 |
| Case 4 | 0.2 | severus_4 | BND | 0 | ENSG00000119772 | *DNMT3A* | chr2 | 25286681 | chr11 | 118488607 | -- | 4 |
| Case 4 | 0.2 | severus_3 | BND | 0 | ENSG00000172493 | *AFF1* | chr4 | 87078149 | chr2 | 25286660 | ++ | 4 |
| Case 4 | 0.2 | severus_4 | BND | 0 | ENSG00000118058 | *KMT2A* | chr11 | 118488607 | chr2 | 25286681 | -- | 4 |
| Case 4 | 0.1 | severus_0 | BND | 0 | ENSG00000119772 | *DNMT3A* | chr2 | 25286660 | chr4 | 87078149 | ++ | 3 |
| Case 4 | 0.1 | severus_0 | BND | 0 | ENSG00000172493 | *AFF1* | chr4 | 87078149 | chr2 | 25286660 | ++ | 3 |

**Supplemental Table 3:** *In silico* limit of detection modeling for paired tumor/normal samples. Pct – percent; SV – structural variant; Chr1 – first chromosome involved in the rearrangement; Chr2 – second chromosome involved in the rearrangement; Supp – supporting.
