## Supplemental Table 6 for "Successful classification of clinical pediatric leukemia genetic subtypes via structural variant detection using HiFi long-read sequencing"

| **Pediatric Leukemia Type** | **Gene** | **HGNC ID** |
| --- | --- | --- |
| Precursor B-cell Acute Lymphoblastic Leukemia | *DUX4* | 50800 |
|  | *ABL1* | 76 |
|  | *BCR* | 1014 |
|  | *EP300* | 3373 |
|  | *ETV6* | 3495 |
|  | *HLF* | 4977 |
|  | *IGH* | 5477 |
|  | *IL3* | 6011 |
|  | *KMT2A* | 7132 |
|  | *MEF2D* | 6997 |
|  | *MYC* | 7553 |
|  | *NUTM1* | 29919 |
|  | *PAX5* | 8619 |
|  | *PBX1* | 8632 |
|  | *RUNX1* | 10471 |
|  | *TCF3* | 11633 |
|  | *ZNF384* | 11955 |
| Acute Myeloid Leukemia | *PML* | 9113 |
|  | *ABL1* | 76 |
|  | *BCR* | 1014 |
|  | *CBFA2T3* | 1537 |
|  | *CBFB* | 1539 |
|  | *CREBBP* | 2348 |
|  | *DEK* | 2768 |
|  | *ERG* | 3446 |
|  | *ETV6* | 3495 |
|  | *FUS* | 4010 |
|  | *GLIS2* | 29450 |
|  | *KAT6A* | 13013 |
|  | *KMT2A* | 7132 |
|  | *MECOM* | 3498 |
|  | *MLF1* | 7125 |
|  | *MNX1* | 4979 |
|  | *MRTFA* | 14334 |
|  | *MYH11* | 7569 |
|  | *NPM1* | 7910 |
|  | *NUP214* | 8064 |
|  | *NUP98* | 8068 |
|  | *RARA* | 9864 |
|  | *RBM15* | 14959 |
|  | *RUNX1* | 10471 |
|  | *RUNX1T1* | 1535 |

**Supplemental Table 6:** Genes utilized for the tumor-only Severus breakend analysis.
